# The impact of non-pharmaceutical interventions on SARS-CoV-2 transmission across 130 countries and territories

**DOI:** 10.1101/2020.08.11.20172643

**Authors:** Yang Liu, Christian Morgenstern, James Kelly, Rachel Lowe, CMMID COVID-19 Working Group, Mark Jit

## Abstract

**Introduction:** Non-pharmaceutical interventions (NPIs) are used to reduce transmission of SARS coronavirus 2 (SARS-CoV-2) that causes coronavirus disease 2019 (COVID-19). However, empirical evidence of the effectiveness of specific NPIs has been inconsistent. We assessed the effectiveness of NPIs around internal containment and closure, international travel restrictions, economic measures, and health system actions on SARS-CoV-2 transmission in 130 countries and territories.

**Methods:** We used panel (longitudinal) regression to estimate the effectiveness of 13 categories of NPIs in reducing SARS-CoV-2 transmission with data from January - June 2020. First, we examined the temporal association between NPIs using hierarchical cluster analyses. We then regressed the time-varying reproduction number (*R_t_*) of COVID-19 against different NPIs. We examined different model specifications to account for the temporal lag between NPIs and changes in *R_t_*, levels of NPI intensity, time-varying changes in NPI effect and variable selection criteria. Results were interpreted taking into account both the range of model specifications and temporal clustering of NPIs.

**Results:** There was strong evidence for an association between two NPIs (school closure, internal movement restrictions) and reduced *R_t_*. Another three NPIs (workplace closure, income support and debt/contract relief) had strong evidence of effectiveness when ignoring their level of intensity, while two NPIs (public events cancellation, restriction on gatherings) had strong evidence of their effectiveness only when evaluating their implementation at maximum capacity (e.g., restrictions on 1000+ people gathering were not effective, restrictions on <10 people gathering was). Evidence supporting the effectiveness of the remaining NPIs (stay-at-home requirements, public information campaigns, public transport closure, international travel controls, testing, contact tracing) was inconsistent and inconclusive. We found temporal clustering between many of the NPIs.

**Conclusion:** Understanding the impact that specific NPIs have had on SARS-CoV-2 transmission is complicated by temporal clustering, time-dependent variation in effects and differences in NPI intensity. However, the effectiveness of school closure and internal movement restrictions appears robust across different model specifications taking into account these effects, with some evidence that other NPIs may also be effective under particular conditions. This provides empirical evidence for the potential effectiveness of many although not all the actions policy-makers are taking to respond to the COVID-19 pandemic.

## Introduction

Coronavirus disease 2019 (COVID-19) is an infectious disease caused by severe acute respiratory syndrome coronavirus 2 (SARS-CoV-2). The virus is easily transmittable between humans, with a basic reproduction number around 2-4 depending on the setting (1,2). To date, no vaccine or highly effective pharmaceutical treatment exists against COVID-19. Countries have used a range of non-pharmaceutical interventions (NPIs) such as testing suspected cases followed by isolation of confirmed cases and quarantine of their contacts, physical distancing measures such as schools and workplaces closures, income support for households affected by COVID-19 and associated interventions, as well as domestic and international travel restrictions (3). These interventions aim to prevent infection introduction, contain outbreaks, and reduce peak epidemic size so that healthcare systems do not become overwhelmed. However, these interventions come at a cost. Testing and contact tracing require laboratory and public health resources to be successful at scale, government subsidies affect national budgets, while physical distancing interferes with economic activities (4). Hence, the psychological, social, and economic cost of interventions needs to be balanced against the potential effectiveness in reducing SARS-CoV-2 spread.

Modelling studies suggest that travel restrictions (5,6), contact tracing and quarantine (7,8) and physical distancing (9,10) may delay SARS-CoV-2 spread. However, the effectiveness of such interventions depends on factors such as societal compliance (e.g., the extent to which people reduce their daily contacts following government restrictions) that are difficult to prospectively measure. Empirical evidence about the effectiveness of specific policy interventions has been limited (11–13). While several countries have seen disease incidence peak and fall (14), ascribing changes in transmission to particular interventions is difficult since countries tend to impose combinations of policy changes at different levels of stringency in close temporal sequence.

Several global databases of COVID-19-related policy interventions have been compiled (15). Here, we used the regularly updated Oxford COVID-19 Government Response Tracker (OxCGRT) (3) and conducted panel analysis to understand the association between policy interventions and time-varying reproduction numbers (*R_t_*), a measure of the rate of transmission of an infectious disease in a population. We also explore whether this relationship is modulated by definitions of policy interventions, temporal lags, and population characteristics in different countries.

## Methods

### Data on NPIs and *R_t_*

Data on COVID-19-related NPI intensity from 1 January to 22 June 2020 was extracted on 5 July 2020 from version 5 of OxCGRT (3), based on the codebook version 2.2 (22 April 2020). This version contains publicly available information from 178 countries and territories on 18 NPI categories. We further divided these countries and territories into seven regions according to the World Bank classification (16). Note that these 18 NPI types are relatively generalised, thus more detailed policy interventions (e.g., facial covering mandate) is not considered in this study.

From this database, we removed (i) “miscellaneous” policies as they contained no data at the time of our data extraction; (ii) “giving international support” and “investment in vaccines” policies as they did not on face validity have a causal pathway to influence local SARS-CoV-2 transmission within the timescale of the analysis; (iii) “fiscal measures” and “emergency investment in healthcare” policies as both the start and the duration of their effect is often unclear (e.g., the announcement of an investment may be implemented weeks later; funding that is allocated may be spent over a long time); (iv) data after 22 June 2020 because >10% of countries and territories have missing data after this date (see Appendix 1). Missing data fields on or before 22 June 2020 were imputed by (a) carrying forward or backwards the next or last non-missing observation when missingness occur at the two tails of the time-series, or (b) linearly interpolating using non-missing observations when missingness does not occur at the two tails of the time series. We divided the remaining 13 policy interventions into four policy groups roughly corresponding to the original database (Table 1).

**Table 1.** Thirteen types of NPIs from OxCGRT, their general categorisations, and the coding schema used in our analysis to quantify their intensity.

| NPI groups | Specific NPIs | Coding schema |
| --- | --- | --- |
| Internal containment and closure | School closure; workplace closure; cancellation of public events; limits on gathering sizes; closure of public transport; stay-at-home requirement; internal movement restriction. | *Any effort scenario:*<br>NPIs are binary variables, considered “present” as long as any (non-zero) effort is made.<br>*Maximum effort scenario:*<br>NPIs are binary variables, considered “present” only if the maximum effort is made.<br>E.g., An intervention X has levels 0-3. A record at level 2 is converted to 1 under *any* and 0 under *maximum scenarios*. |
| International travel restrictions | International movement restriction. |  |
| Economic policies | Income support; debt/ contract relief for households |  |
| Health systems policies | Public information campaign; testing policy; contact tracing |  |

Most NPIs in the database are measured on ordinal scales that capture intensity (e.g. 0 - no contact tracing; 1 - limited contact tracing; 2 - comprehensive contact tracing). Since the intervals between categories are not necessarily equally spaced, we converted NPIs history into binary variables under: (i) *any effort scenario*: all zero records were converted to 1, non-zero records were converted to 0; and (ii) *maximum effort scenario*: all non-maximum records were converted to 0, all records at maximum levels were converted to 1 (see Table 1).

Transmission of SARS-CoV-2 is routinely measured using the time-varying reproduction number (*R_t_*), a metric which represents the mean number of secondary cases that one index case will infect. We used the median *R_t_* estimates available through EpiForecast. The estimation process is based on reported incidence while accounting for a range of uncertainties surrounding the incubation period, the delays between symptom onset and reporting (17). The underlying method has been detailed in Cori et al (18). In short, the transmission rate of an infectious disease is approximated by the ratio between new infections at time t and the infectious individuals at time t - *w* where *w* is the associated time window. In EpiForecast, a weekly time window is used. This measure is expected to fall when effective NPIs reduce the rate of SARS-CoV-2 transmission. Since the effects of some NPIs may take time to become evident, we explored a range of temporal lag effects between NPI implementation and *R_t_* changes.

Between 1 January and 22 June 2020, data on NPIs and *R_t_* are simultaneously available for 130 countries and territories, all of which are used in the panel analysis described below.

### Understanding the Temporal Patterns

The effect of an NPI may vary as a result of the evolving epidemic dynamics (e.g., decreasing number of susceptibles) or time-varying factors such as public compliance (e.g., the proportion of shoppers wearing facial covering after government mandate). To examine this effect, we split up the time series of NPIs and *R_t_* values into two parts: before and after peak NPI stringency. This can be considered a sensitivity analysis to examine the robustness of NPIs’ effectiveness in reducing COVID-19 transmission across time.

We used OxCGRT’s stringency index (SI), a combined metric of several behaviour-related NPI measures, as a proxy to the overall constraint on people’s day-to-day life. We then fitted a Gaussian generalised additive model (GAM) with cubic splines, using SI as the response variable and date as the sole explanatory variable for each World Bank region (i.e., the predicted regional SI is informed by all stringency index time-series within it). The peak of the predicted SI splines for each region was then examined to derive an average peak across all the regions. We then constructed two time-series: (i) the *full* time series, and (ii) the *truncated* time series up to the time of peak SI.

We also examined temporal clustering among different NPIs. This is important because the impact of two NPIs that are highly temporally clustered may not be independently identifiable. During variable selection in panel regression, one of the two NPIs may be removed due to multicollinearity, but this does not mean for certain that the NPI removed is not associated with the observed effects. To investigate the potential temporal clustering, we conducted hierarchical cluster analysis using Ward’s method (19) under the *any effort scenario* and the *maximum effort scenario*. We then used multi-scale bootstrapping (n = 10,000) to test the statistical significance of the identified clusters, defined using approximate unbiased p-values less than 0.05 (20).

### Panel analyses

We used panel (or longitudinal) regression to study the association between NPI intensity and *R*_t_, treating the time-series of NPI intensity and *R_t_* in each country as observations of an individual in a panel. A linear fixed effects model:

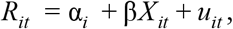

where *R_it_* is the time-varying reproduction number of location *i* at time *t*; α*_i_* is a location-specific intercept (assumed to remain constant over the timescale of the analysis); β*X_it_* represents the 13 NPIs and their corresponding coefficients; and *u_it_* is the error term. The decision of using a fixed-effect model with individual intercept (as opposed to a random effect model) is based on the results of Hausman test (21).

We investigated the appropriate temporal lag between NPI intensity and *R_t_*. To do this, we calculated the deviance (logarithm of the sum of squared residuals divided by the number of data points) assuming errors are normally distributed for temporal lags of 1 to 21 days. Smaller deviances indicate temporal lags that give better fits to data. A temporal lag of *k* days regresses the *R_t_* on a particular day with NPIs implemented *k* days before (i.e.*X_i,t_*_−_*_k_*). This analysis was carried out on both the regional and the global level. Data from North America and South Asia were excluded from region-specific temporal lag analyses due to small sample sizes.

Stepwise backwards variable selection based on Akaike or Bayesian Information Criterion (AIC or BIC) was then used to choose the most parsimonious model. Beginning with the full model (13 independent variables, one for each NPI), independent variables were removed one at a time sequentially.

### Statistical Interpretation

For both the *any effort* and the *maximum effort scenarios*, we examined a range of model specifications consisting of (i) different variable selection criteria: AIC and BIC, (ii) different temporal lags between the timing of NPIs and changes in *R_t_* (selected based on deviance from the analysis of temporal patterns discussed above), and (iii) different time series lengths: one ending on 22 June 2020 and the other truncated to 13 April 2020, when NPI intensity peaked (on average). We then defined categories of ‘evidence strength’ behind each association according to Table 2. Allocating each NPI to an evidence category was done independently by two authors (YL and MJ), with differences resolved by discussion.

**Table 2.** Expert interpretation of evidence behind the statistical associations of each NPI with reductions in *R_t_*.

| Evidence Strength | Effect estimates | Temporal Cluster |
| --- | --- | --- |
| Strong | Selected and significant with intended effect signs (i.e. negative) regardless of model specifications (i.e., variable selection criteria, temporal lags, and time-series lengths). | Not in a temporal cluster with any NPI with significantly positive effect estimates. |
| Moderate | Selected and significant with intended effect sign (i.e negative) in two of three model specification dimensions (i.e., variable selection criteria, temporal lags, and time-series lengths), and non-selected or non-significant in the remaining dimension.* |  |
| Weak | Not strong or moderate |  |

\* For the moderate category, all the model specifications that were non-significant or non-selected were examined to see if they had a value in common across one of the three criteria e.g. all of them had a lag of 10 days.

### Software

All analyses were conducted using R version 4.0.0 (22), with packages ̀plm̀ and ̀pvclust̀ (23,24). Code is available at https://github.com/yangclaraliu/COVID19 NPIs vs Rt.

## Results

### Trends in NPI intensity

Temporal trends in COVID-19-related NPI intensity measured using the OxCGRT SI are relatively consistent across regions (Figure 1). Following the initial NPIs in China, almost all regions experienced an initial increase in policy stringency in early February 2020. The East Asia & Pacific region had the highest SI up to mid-March, but by April had the lowest SI. From March, other regions registered rapid increases in their stringency index. The stringency index peaked in mid-April for all regions, and so 13 April 2020 was used as the time of peak NPI intensity (see Appendix 2). All regions and nearly all countries had a higher stringency index in June compared to February.

**Figure 1.**
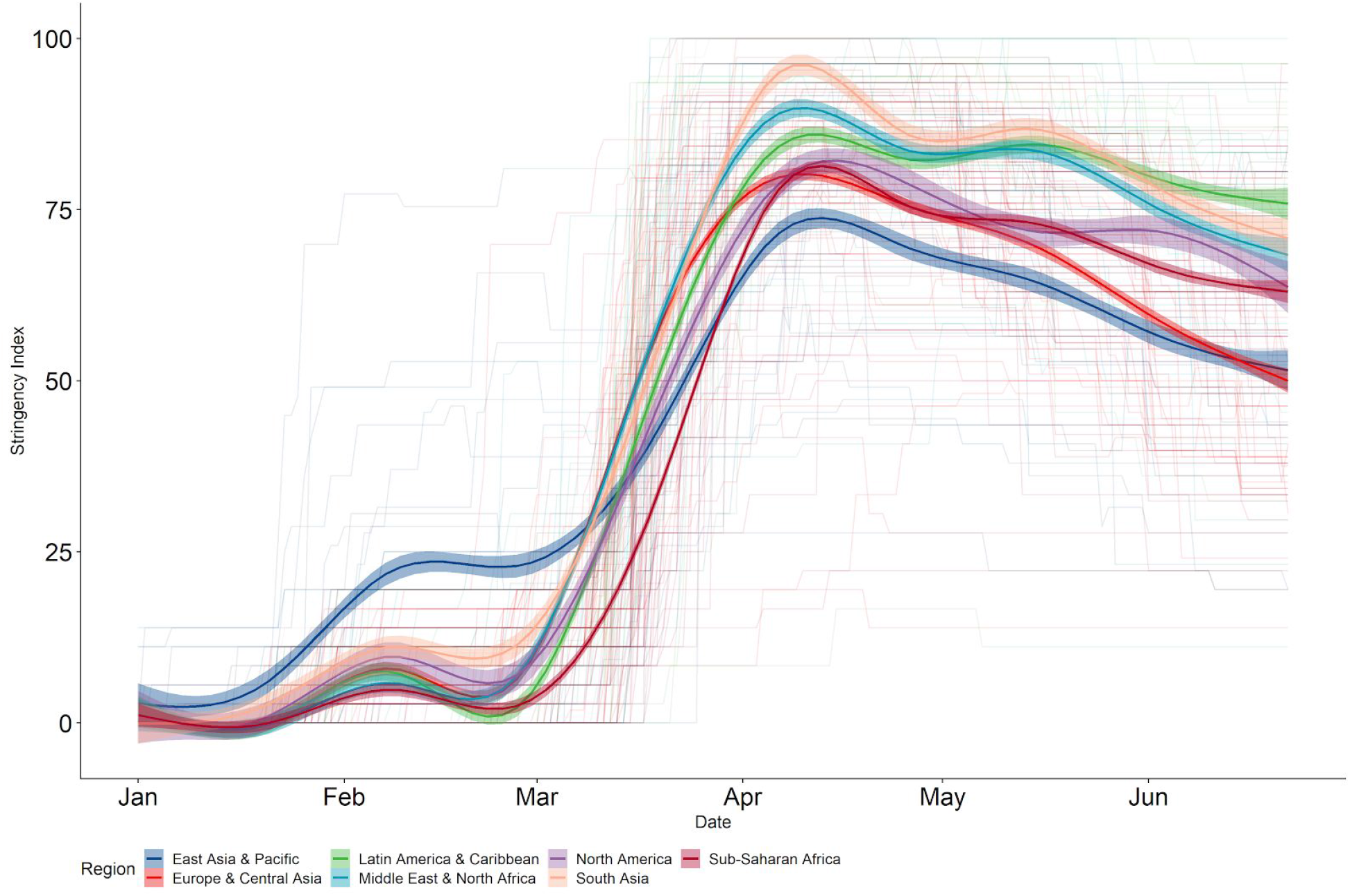
Temporal changes in NPI stringency index (range = 0-100) by region. Countries with available data are assigned corresponding geographical regions based on the World Bank classification scheme. Although Asia increased policy stringency first (observed in Feb), the highest stringency achieved was the lowest (observed in April). Countries become less stringent in terms of COVID-19 response at approximately the same time worldwide.

Figure 2 shows how the intensity of specific NPI groups varies in each region relative to the time of peak intensity. Under both *any* and *maximum effort scenarios*, ‘Health System Policies’ was the first NPI group to increase across all regions. It was also the most commonly used NPI group. This was followed by ‘Internal Containment and Closures’ and then ‘Economic Policies’, although ‘International Travel Restrictions’ sometimes came before ‘Internal Containment and Closures’. NPI intensity increased only as the first case was detected in each region, except for Sub-Saharan Africa where many countries took action before the first case. While SIs have decreased across all regions (Figure 1), this is not very apparent in most NPI groups apart from International Travel Restrictions (Figure 2).

**Figure 2.**
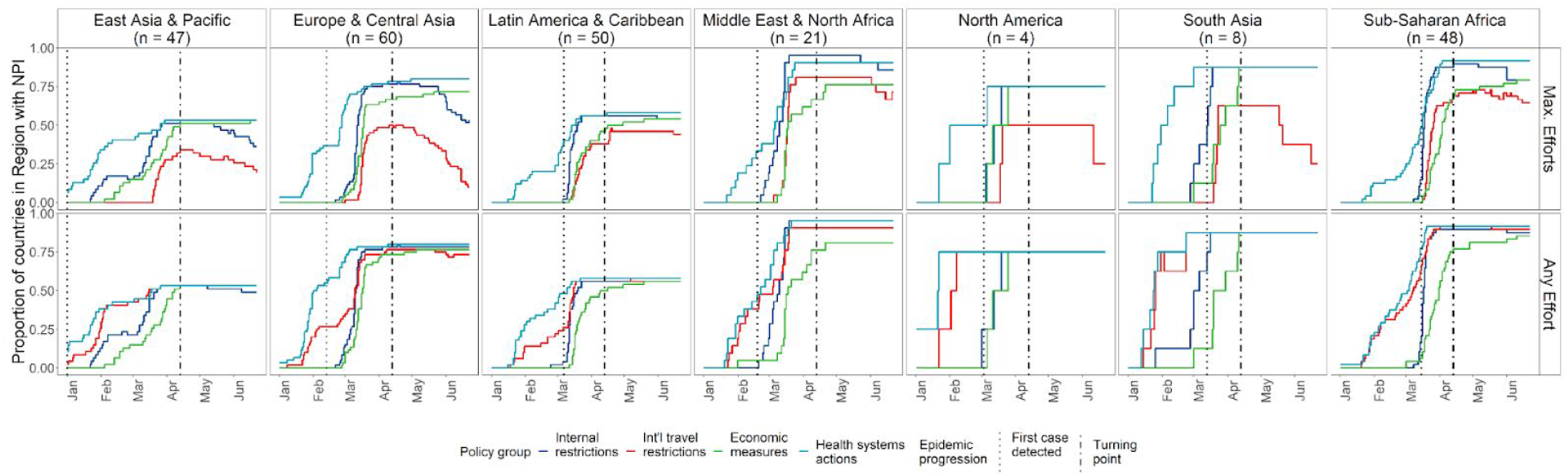
The proportions of countries implementing NPIs in each group by region. Colours indicate different NPI groups, as defined in Table 1; *n* values represent the total number of countries and territories in each region per definitions of the World Bank; the y-axis shows the proportion of countries and regions implementing given NPIs, e.g., proportion of one means everyone within the region are implementing the NPI; turning point is selected based on the region-specific peaks of predicted Stringency Index, shown in Figure 1.

Hierarchical cluster analysis shows that, given the *any effort scenario*, all the NPIs are contained in two significant temporal clusters (Figure 3). These temporal clusters align well with the broad categorisations defined in the OxCGRT, i.e. countries tend to start implementing the same categories of NPI simultaneously. However, under the *maximum effort scenario*, there are three significant temporal clusters and several NPIs are not in any cluster, i.e. countries reach their maximum level of intensity for NPIs at very different times.

**Figure 3.**
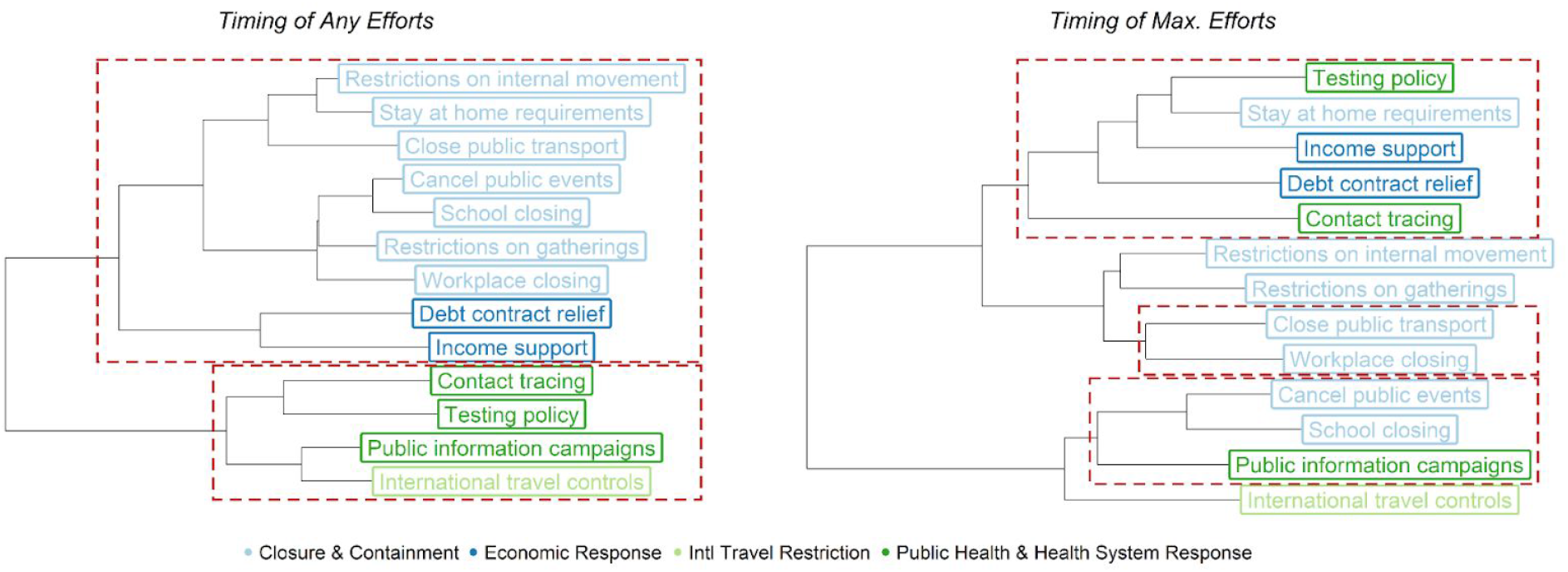
Hierarchical cluster analysis of NPIs time-series by scenario. Blues and greens are used to describe the corresponding NPI groups; red boxes indicate statistically significant temporal clusters. Hierarchical clustering was carried out using Ward’s method;the statistical significance of temporal clusters identified was obtained via bootstrapping.

### Panel analyses

We examined the goodness-of-fit (based on deviance) of the panel regression model in all scenarios both at the regional and global level to identify the most appropriate temporal lag (Appendix 3). For both the *full* and *truncated* time series (ending on 22 June and 13 April 2020, respectively), we identified temporal lags to be longest in East Asia & Pacific (between 5 and 10 days), followed by Europe & Central Asia (approximately 5 days), and the shortest in Latin America & Caribean (below 5 days) (see also A3). The results from the Middle East & North Africa and Sub-Saharan were not consistent, implying dependency on the scenario used, and there was not a clear indication of the most appropriate temporal lag when countries were all combined in a global analysis. Due to the observed heterogeneity in the temporal lags, we examined three different lag values (1, 5, and 10 days) in the regression analyses for both *full* and *truncated* time-series.

The NPIs in the models selected based on AIC and BIC are shown in Figure 4. Under the *any effort scenario*, the most consistently excluded NPIs are contact tracing, restrictions on gatherings, and international travel restrictions. Public information campaigns and testing policies were excluded using the *truncated* but not the *full* time-series; stay at home requirements was excluded using the *full* but not the *truncated* time-series. Under the *maximum effort scenario*, the most consistently excluded NPIs are contact tracing, international travel restrictions, and closure of public transportation. Public information campaigns were excluded by one model using the *truncated* time series but were always included by models using the *full* time-series. NPIs may be excluded from models either because (i) they do not affect *R_t_* or (ii) their effects were fully captured by other NPIs in the same temporal clusters and thus they were removed by the variable selection process.

**Figure 4.**
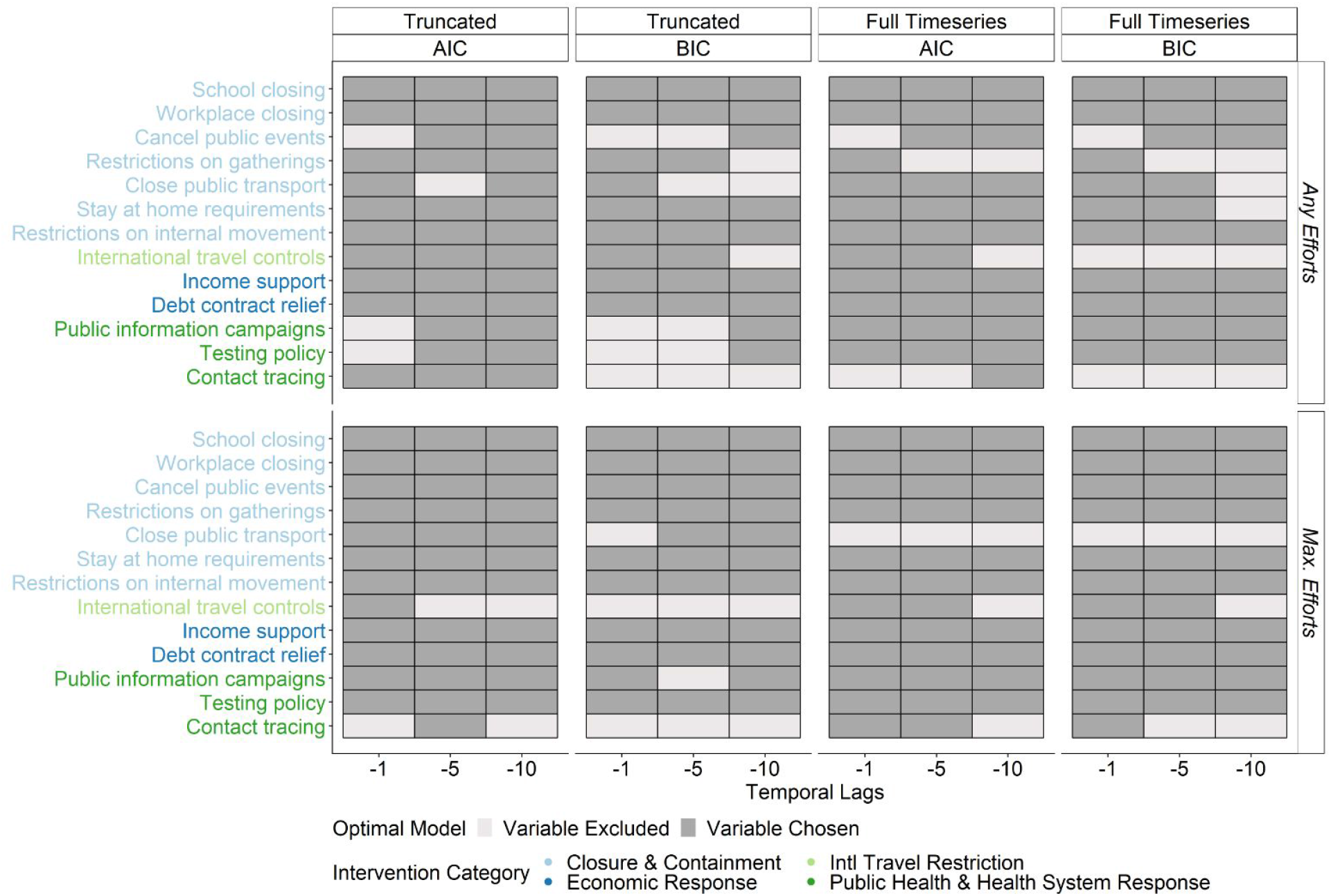
Variable selection results. Optimal models are based on a backward step-process using AIC/BIC. NPIs are colour coded based on their respective NPI categories.

Effect size estimates for the selected models in Figure 4 are shown in Figure 5. A few NPIs have significantly positive effects, indicating that they are associated with increased *R_t_*. While it is not inconceivable that some NPIs may be transiently associated with increased *R_t_* (e.g. increased testing efforts may be associated with increased *R_t_* because they result in better detection of COVID-19 cases), variables with positive effects are likely capturing residual non-random errors for other NPIs in the same temporal cluster. Hence they are likely biasing effect size estimates of other temporal cluster members, likely away from the null hypotheses. Hence, NPIs that are temporally related need to be interpreted within the context of the respective clusters rather than as individual measures (see also Figure 3).

**Figure 5.**
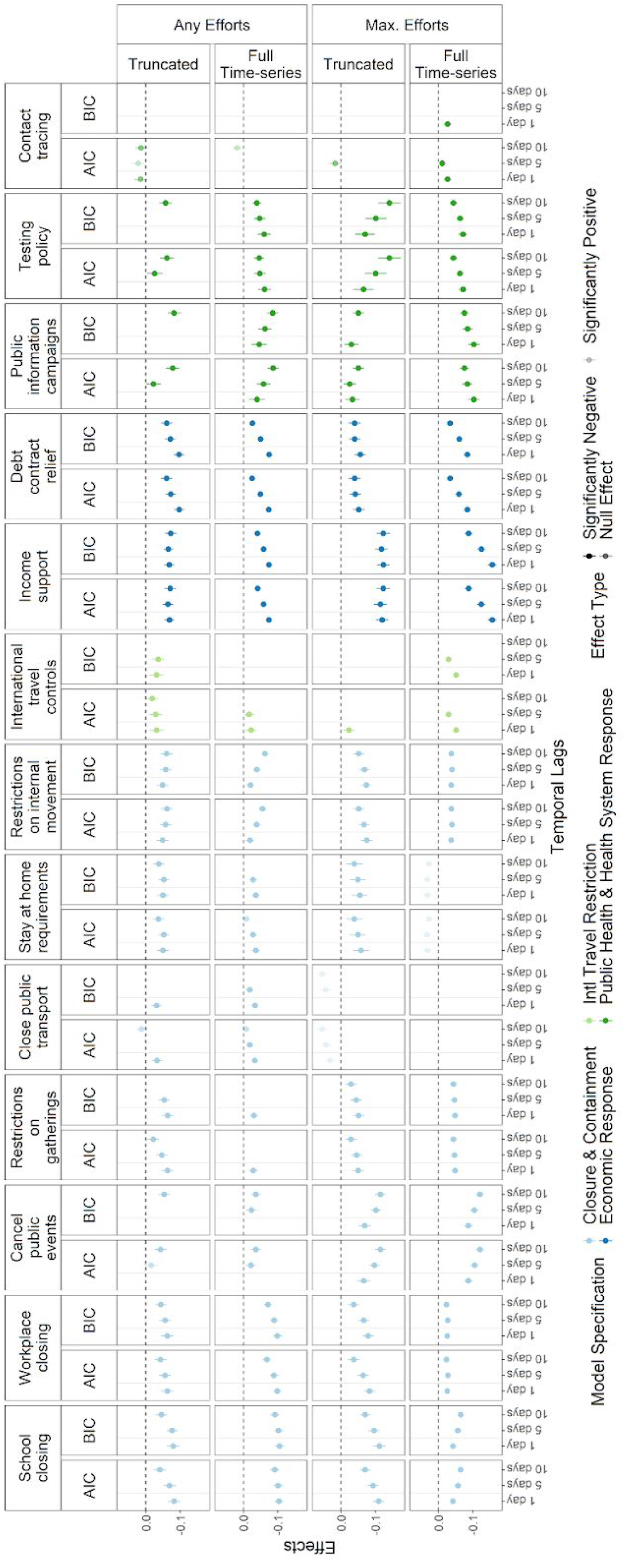
Effect sizes for each NPI from the selected models. Points and lines indicate mean and 95% confidence intervals.

### Interpretation

Of the 13 NPIs in the OxCGRT, we found strong evidence for the association between two of them (school closure, internal movement restrictions) and *R_t_*, under both *any effort* and *maximum effort* scenarios. Another three NPIs (workplace closure, income support and debt/contract relief) had strong evidence for the association under the *any effort* scenario only, meaning that the reductions in *R_t_* were associated with the initiation of these interventions, with no evidence of an effect as they were intensified. Two other NPIs (public events cancellation, restriction on gathering) had strong evidence under the *maximum effort* scenario only, meaning that evidence for a reduction in *R_t_* was only seen when they reached their maximum intensity.

It is probable that in some cases, interventions occurring with certain sequential order may make it more or less likely candidates to capture the effects of other NPIs. For example, the method used in Abbott et al. (17), which relies on back sampling to convert onset to confirmation timing, may move incidence back in history more than reality. This shifts the time series of *R_t_* and NPIs, increasing the probability that NPIs occurring later in the COVID-19 to be associated with reduced *R_t_*. Thus, we verify NPIs supported by strong statistical evidence by checking their sequential orders in COVID-19 response strategies. Most of these NPIs were not implemented particularly early or late (Appendix 4). Complete school closure and mandatory public events cancellations are moderately skewed earlier, indicating that they tend to occur first. Some (non-maximum) levels of income support and debt/ contract relief are skewed later, making it possible their observed effects are statistical artefacts.

Evidence for the rest of the NPIs was decidedly mixed. Stay-at-home requirements had moderate evidence under the *any effort* scenario, while public information campaigns had moderate evidence under the *maximum effort* scenario. Among all NPIs, some (non-maximum) levels of stay-at-home requirements tend to occur later in the overall COVID-19 response strategy. Most public places people would like to visit have already closed - thus making the relevant effects minimum. The remaining four (public transport closure, international travel controls, testing, contact tracing) had only weak evidence for an association with *R_t_*. Detailed interpretation on the statistics, through which these conclusions were reached, is presented in Appendix 3. Similar methods were applied to the original raw data, without converting to *any* or *maximum effort scenarios*. However, as no statistical conclusion can be reasonably drawn, we only show the results in Appendix 5.

We observed variability in effect estimates due to differences in terms of time-series and temporal lags used (Figure 5). For example, the effect sizes of internal movement restrictions are smaller using the truncated time-series compared to using the full time-series. This suggests that the general adherence to movement restrictions may have decreased over time. However, this variability may also be explained by the fact that full-time series include more observations. The effect sizes of public events cancellation is higher for longer temporal lags - indicating its impacts on *R_t_* may take time to be noticeable. These hypotheses need further validation using empirical evidence.

## Discussion

Our study used panel regression to examine the temporal association between NPIs that countries introduced in response to the COVID-19 pandemic, and its rate of transmission in populations, represented by *R*_t_. We explored the influence of different model specifications on the association: (i) whether NPIs could be at the minimum intensity or had to take their maximum intensity values to effectively reduce *R_t_*, (ii) whether the effect of NPIs was different at the start of the pandemic compared to after NPI intensity had peaked globally, (iii) different temporal lags between NPIs and their effects and (iv) different criteria for selecting NPIs as explanatory variables.

We found the strength of evidence behind an association between NPIs and *R_t_* depended on these model specifications. Only two NPIs (school closure, internal movement restrictions) showing unequivocal evidence of being associated with a decrease in *R_t_* regardless of the assumptions made. Whether schools should stay closed has drawn debates. Keeping schools closed could potentially hurt children’s educational development and general wellbeing. Resuming schools, on the other hand, may increase COVID-19 transmission risks for both students and teachers. Our findings are consistent with much existing literature - although school closures cannot single-handedly suppress an outbreak, they are generally effective in terms of reducing transmission (25,26).

We found evidence that internal movement restrictions reduced *R_t_*, but no evidence of a similar effect for international travel restrictions. The latter is consistent with Russell et al., which shows international movement restrictions have limited impacts on the epidemic dynamics of COVID-19 for most countries (27). This difference may be explained by the different decision thresholds (e.g., real-time expected *R_t_* at which governments implement travel restrictions). It may relate to the types of movement interrupted - internal movement restrictions interrupt trips of all lengths whereas international movement restrictions may have disrupted more longer trips. Last but not least, these NPIs were likely used in different epidemic contexts - internal movement restrictions used more often to prevent outbreaks from escalating whereas international travel restrictions to prevent infection introduction (28). The latter was not necessarily captured by *R_t_*, which could only be estimated in settings with existing COVID-19 outbreaks (i.e., after introduction).

There are differences in the strength and direction of the effects of some NPIs (such as public transport closure and stay-at-home requirements) depending on whether the whole time series of data was used, or only data up to the date of peak NPI stringency (13 April 2020). This may indicate that these NPIs might have different effects at the start of the pandemic compared to later on, so when the NPIs were removed (likely after the peak), *R_t_* did not return to its original level before the introduction of the NPIs.

The best-fitting models also support a considerable delay between NPIs and their effect on transmission. This delay is about a week on average but differs widely between regions. It could reflect delays between policies being put in place and actual behaviour change. It could also reflect delays in reporting; although these are explicitly accounted for in the *R_t_* estimation in EpiForecasts - the same onset-to-delay distribution is applied in all countries (29), and hence may not reflect differences between settings. Delays of up to 3 weeks between policy changes and changes in reported cases have been documented (30).

We were not able to uncover evidence that supports the effectiveness of contact tracing and testing policies. This may be explained by the fundamental differences in nature to these NPIs - both contact tracing and testing policies, besides interrupting onward transmission, could lead to more cases being reported. While calculating the *R_t_*, EpiForecasts does not explicitly account for changes in reporting rate (17). The estimated effects are the sum of opposing effects. Another potential explanation is the way NPIs are reported in the OxCGRT, which largely relies on publicly available data sources, such as news articles. Contact tracing and test policies are both classic public health intervention tools and have minimal impacts on those who are not potentially infected. Thus, they may be less likely to receive media coverage, compared to more interruptive NPIs such as workplace closures.

Many other papers have explored the impact of physical distancing measures on SARS-CoV-2 transmission. Prospective mechanistic transmission models have explicitly modelled contacts relevant to viral transmission between individuals in different subgroups (e.g. ages), as well as the impact that NPIs may have on these contacts. Such studies mainly use data from a single location only such as Wuhan (9), Hong Kong (31), the United States (32) and the United Kingdom (26). They suggest that physical distancing interventions can have a large impact on transmission. While the impact of income-related interventions have been less well studied, country reports suggest that they often play an important role in ensuring adherence to distancing measures (33).

Another group of studies have used empirical data to retrospectively examine whether NPIs have been effective in reducing transmission, using either statistical methods or mechanistic epidemiological models. Many such studies look at single interventions such as travel restrictions (34) or “lockdowns” (35,36). Hence they are less useful to policy-makers wanting to establish which of a basket of NPIs are most effective.

Only a small number have looked at multiple interventions across multiple countries. These relate NPIs from databases to proxies of transmission such as *R_t_* estimated from cases and/or deaths (11,12), or the rate of change in cases directly (13,37,38). Our work demonstrates the major challenges that all such studies (including ours) face - NPI introductions are highly temporally correlated in time, so it is difficult to independently identify the effect of each NPIs.

Our study extends previous work to address this problem in several ways. Firstly, we use data across a larger number of countries and territories and longer time series (January - June 2020), enhancing the power to detect independent effects even when there is partial collinearity. Second, instead of assuming that all NPIs tested have an effect like previous work, we conduct variable selection to identify only those NPIs that are retained in parsimonious models. Third, we conduct cluster analysis to explicitly identify temporal correlations, and use this in our interpretation of the strength of evidence behind each intervention. Fourth, we have conducted sensitivity analyses across a range of model specifications around the variable selection criteria, temporal lag between NPIs and change in transmission, temporal truncation, as well as the way NPI intensity is coded.

Nonetheless, our study also has several limitations. First, besides the information bias in the NPIs database discussed above, the coding scheme may also introduce potential bias. NPIs coded as “comprehensive contact tracing for all identified cases” may have different implications in different countries. Effectiveness of contact tracing in places like Singapore (39) may be masked by seemingly similar but realistic non-comparable contact tracing programs. Second, compared to daily incidence, *R_t_* estimates are much more suitable to compare across countries and thus is used as the metric for COVID-19 transmission in this study. However, these estimates are based on a series of assumptions (e.g., distribution of onset to confirmation delay) that may not always be appropriate. Our current model also does not factor in uncertainty around *R_t_* estimates. Last but not the least, although we examined a wide range of NPIs, we did not include any potential interactions in the current model. Such interaction is a possibility, e.g., more people may comply with workplace closures when receiving income support. Future research should look into these relationships.

## Conclusions

In conclusion, evidence from a panel of 130 countries and territories, provides evidence about the effectiveness of school closure, public events cancellations, and household income support in reducing SARS-CoV-2 transmission. Despite the inherent limitations of observational and ecological data, our study provides the broadest empirical evaluation so far that NPIs can work.

## Data Availability

All data is availability through database described in this manuscript. Code used can be found here [https://github.com/yangclaraliu/COVID19_NPIs_vs_Rt].

## Acknowledgements

CM and JK’s contribution to this work was supported by the Royal Society’s Rapid Assistance in Modelling the Pandemic (RAMP) scheme.

We thank Richard Pebody (WHO European Region Office, Copenhagen), Katelijn Vandemaele (WHO, Geneva) and Helen Johnson (European Centre for Disease Prevention and Control, Stockholm) for helpful comments.

YL and MJ are funded by the National Institute of Health Research (UK)(16/137/109), the Bill & Melinda Gates Foundation (INV-003174), and the European Commission (101003688). This research was partly funded by the National Institute for Health Research (NIHR) (16/137/109) using UK aid from the UK Government to support global health research. The views expressed in this publication are those of the author(s) and not necessarily those of the NIHR or the UK Department of Health and Social Care. This research is partly funded by the Bill & Melinda Gates Foundation (INV-003174). The findings and conclusions in this report are those of the author(s) and do not necessarily represent the official position of the Bill & Melinda Gates Foundation. YL is also supported by the UK Medical Research Council (MC_PC_19065). CM and JK are employed by IPM Informed Portfolio Management. IPM is appreciative of the contributions of its employees above and beyond the scope of their work, their dedication to the community and the world as a whole. The views expressed herein are made in a personal capacity and are not those necessarily made, sponsored, affiliated or endorsed by IPM. RL is a UK Royal Society Dorothy Hodgkin Fellow.

Funding information for the Centre for Mathematical Modelling of Infectious Disease COVID-19 Working Group: James Munday (Wellcome Trust: 210758/Z/18/Z); Hamish Gibbs (UK DHSC/UK Aid/NIHR: ITCRZ 03010); Carl A B Pearson (BMGF: NTD Modelling Consortium OPP1184344, DFID/Wellcome Trust: 221303/Z/20/Z); Kiesha Prem (BMGF: INV-003174, European Commission: 101003688); Quentin J Leclerc (UK MRC: LID DTP MR/N013638/1); Sophie R Meakin (Wellcome Trust: 210758/Z/18/Z); W John Edmunds (European Commission: 101003688, UK MRC: MC_PC_19065, NIHR: PR-OD-1017-20002); Christopher I Jarvis (Global Challenges Research Fund: ES/P010873/1); Ammy Gimma (Global Challenges Research Fund: ES/P010873/1, UK MRC: MC_PC_19065); Sebastian Funk (Wellcome Trust: 210758/Z/18/Z); Matthew Quaife (ERC Starting Grant: #757699, BMGF: INV-001754); Timothy W Russell (Wellcome Trust: 206250/Z/17/Z); Jon C Emory (ERC Starting Grant: #757699); Sam Abbott (Wellcome Trust: 210758/Z/18/Z); Joel Hellewell (Wellcome Trust: 210758/Z/18/Z); Rein M G J Houben (ERC Starting Grant: #757699); Kathleen O’Reilly (BMGF: OPP1191821); Georgia R Gore-Langton (UK MRC: LID DTP MR/N013638/1); Adam J Kucharski (Wellcome Trust: 206250/Z/17/Z); Megan Auzenbergs (BMGF: OPP1191821); Billy J Quilty (NIHR: 16/137/109, NIHR: 16/136/46); Thibaut Jombart (Global Challenges Research Fund: ES/P010873/1, UK Public Health Rapid Support Team, NIHR: Health Protection Research Unit for Modelling Methodology HPRU-2012-10096, UK MRC: MC_PC_19065); Alicia Rosello (NIHR: PR-OD-1017-20002); Oliver Brady (Wellcome Trust: 206471/Z/17/Z); Kevin van Zandvoort (Elrha R2HC/UK DFID/Wellcome Trust/NIHR, DFID/Wellcome Trust: Epidemic Preparedness Coronavirus research programme 221303/Z/20/Z); James W Rudge (DTRA: HDTRA1-18-1-0051); Akira Endo (Nakajima Foundation, Alan Turing Institute); Kaja Abbas (BMGF: OPP1157270); Fiona Yueqian Sun (NIHR: 16/137/109); Simon R Procter (BMGF: 0PP1180644); Samuel Clifford (Wellcome Trust: 208812/Z/17/Z, UK MRC: MC_PC_19065); Nicholas G. Davies (NIHR: Health Protection Research Unit for Immunisation NIHR200929, UK MRC: MC_PC_19065); Charlie Diamond (NIHR: 16/137/109); Rosanna C Barnard (European Commission: 101003688); Rosalind M Eggo (HDR UK: MR/S003975/1, UK MRC: MC_PC_19065); Emily S Nightingale (BMGF: OPP1183986); David Simons (BBSRC LIDP: BB/M009513/1); Katharine Sherratt (Wellcome Trust: 210758/Z/18/Z); Graham Medley (BMGF: NTD Modelling Consortium OPP1184344); Gwenan M Knight (UK MRC: MR/P014658/1); Stefan Flasche (Wellcome Trust: 208812/Z/17/Z); Nikos I Bosse (Wellcome Trust: 210758/Z/18/Z); Petra Klepac (Royal Society: RP\EA\180004, European Commission: 101003688)

## Technical Appendix

### A1. Missingness in data

The figure below shows the number of countries and regions each day between 1 January - 5 July 2020 with at least one non-missing entry in Oxford COVID-19 Government Response Tracker (3). Vertical solid line indicates the cutoff threshold of 22 June 2020 used in this study.

**Figure S1.**
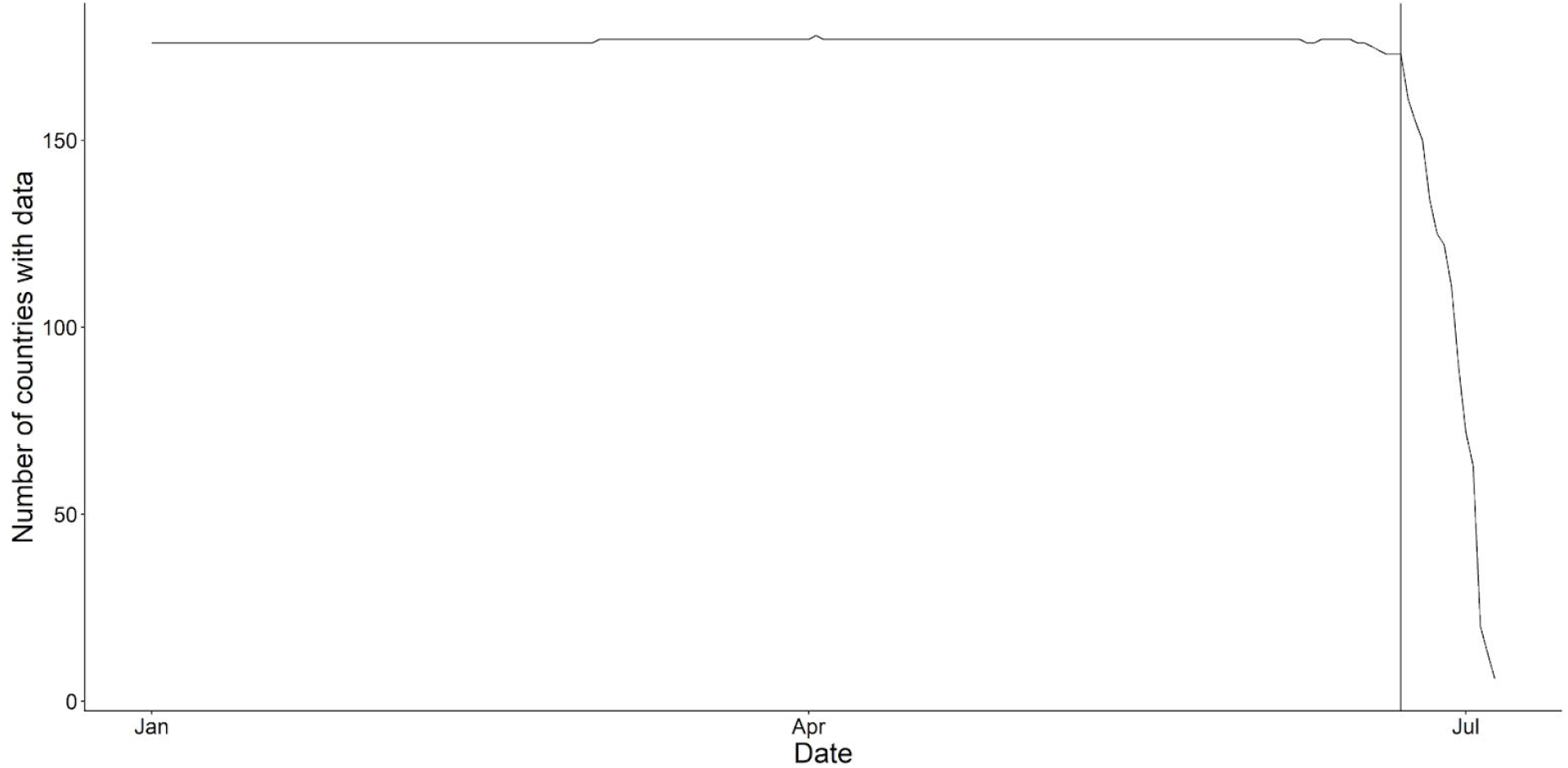
Number of countries and regions with available data in the Oxford COVID-19 Government Response Tracker.

### A2. Peak timing of stringency indices by region

**Table S1.** * Peak date is defined as the date corresponding to the highest predicted stringency index on the regional level based on the general additive models.

| Region | Peak Date |
| --- | --- |
| East Asia & Pacific | 2020-04-13 |
| Europe & Central Asia | 2020-04-11 |
| Latin America & Caribbean | 2020-04-13 |
| Middle East & North Africa | 2020-04-11 |
| North America | 2020-04-15 |
| South Asia | 2020-04-11 |
| Sub-Saharan Africa | 2020-04-13 |

### A3. Temporal Lags

The original Oxford COVID-19 Government Response Tracker contains NPIs information in the format of ordinal categorical variables (3). Take “Restrictions on Gatherings” for example, there are five levels:

0 - No restrictions;
1 - Restrictions on very large gatherings (the limit is above 1000 people)
2 - Restrictions on gatherings between 101-1000 people
3 - Restrictions on gatherings between 11-100 people
4 - Restrictions on gatherings of 10 people or less.

As discussed in the main text, we analysed two key scenarios:

- *Any effort scenario*: NPIs are binary variables, considered “present” as long as any (non-zero) effort is made;
- *Maximum effort scenario*: NPIs are binary variables, considered “present” only if the maximum effort is made.

For a hypothetical country’s, the NPI time-series regarding “Restrictions on Gathering” such as {0, 0, 0, 1, 1, 2, 3, 4, 4} is converted to:

- *Any effort scenario* as {0, 0, 0, 1, 1, 1, 1, 1, 1}
- *Maximum effort scenario* as {0, 0, 0, 0, 0, 0, 0, 1, 1}

An additional scenario - *multilevel effort scenario*, which preserves the original time series without conversion, was originally explored but had to be excluded due to severe temporal clustering and biased effect size estimates. However, here in the appendix we still showed some of these results, showcasing why they have not been included in the main discussion. For more details, please also see Appendix 5.

**Figure S2.**
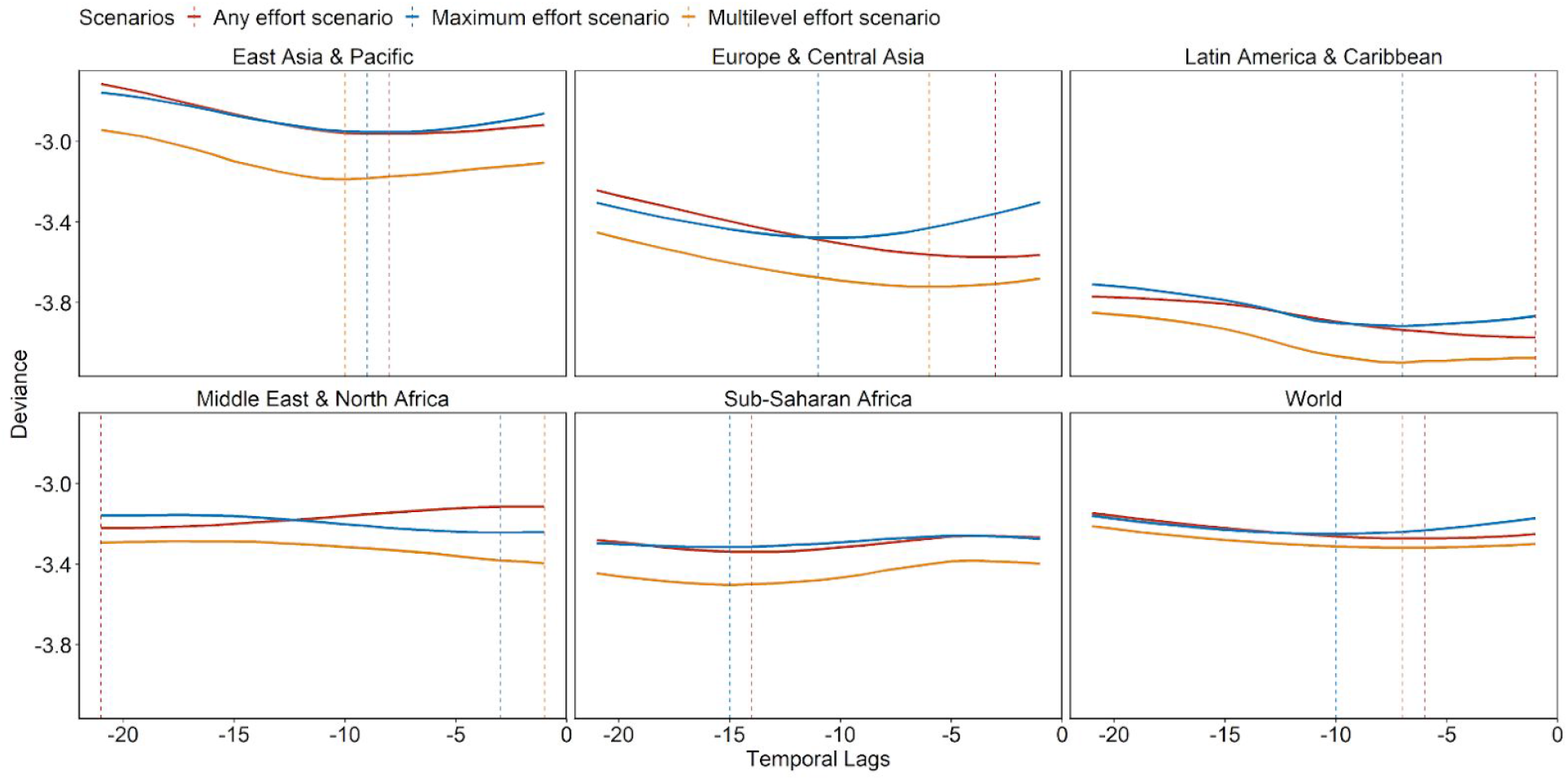
Deviance from panel analyses using different temporal lags between effective reproduction number and policy interventions. Models include all 13 interventions available. Deviance is defined as the logarithm of the sum of squared residuals divided by the number of data points. Full time-series from 1 Jan to 22 June 2020 were used. Dashed vertical lines indicate minimum deviances.

**Figure S3.**
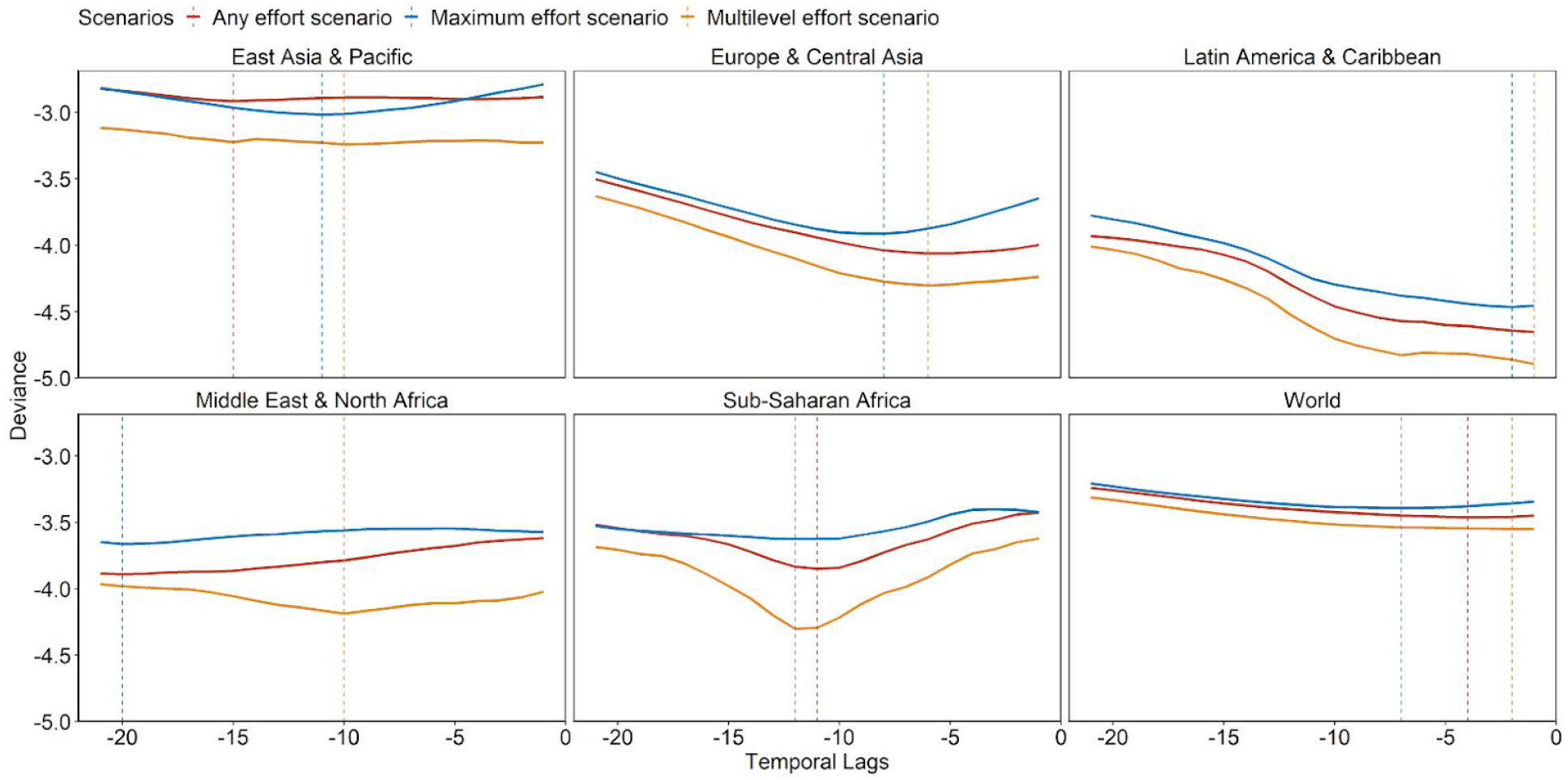
Deviance from panel analyses using different temporal lags between effective reproduction number and policy interventions. Models include all 13 interventions available. Deviance is defined as the logarithm of the sum of squared residuals divided by the number of data points. Truncated time-series from 1 Jan to 13 April 2020 were used. The turnpoint on 13 April 2020 was identified using stringency indices. Dashed vertical lines indicate minimum deviances.

### A4. Interpretation of Panel Analyses Results

**Table S2.** Statistical interpretation worksheets by YL and MJ detailing how these conclusions are reached can be downloaded from [https://docs.google.com/spreadsheets/d/1lZUuxgXc6ZPCx4IA56iYwokuDgU2if9nTySfDwqi50/edit?usp=sharing].

| Non-pharmaceutical Interventions | Any Efforts Scenario | Maximum Efforts Scenario |
| --- | --- | --- |
| School closures | Strong | Strong |
| Workplace Closure | Strong | Weak |
| Public Events Cancellation | Weak | Strong |
| Restriction on gathering | Moderate | Strong |
| Public Transportation Closures | Weak | Weak |
| Stay-at-home requirements | Moderate | Weak |
| Internal Movement Restrictions | Strong | Strong |
| International Travel Controls | Weak | Weak |
| Income Support | Strong | Weak |
| Debt/ Contract Relief | Strong | Weak |
| Public Information Campaign | Weak | Moderate |
| Testing | Weak | Weak |
| Contact tracing | Weak | Weak |

### A4. Implementation Sequence of Non-pharmaceutical Interventions

A new ranking variable was created for each NPI in each scenario. For example, the NPI implemented first is ranked 1. The distributions of the sequential order for each NPI is shown below for both *any* and *maximum efforts* scenarios.

**Figure S4.**
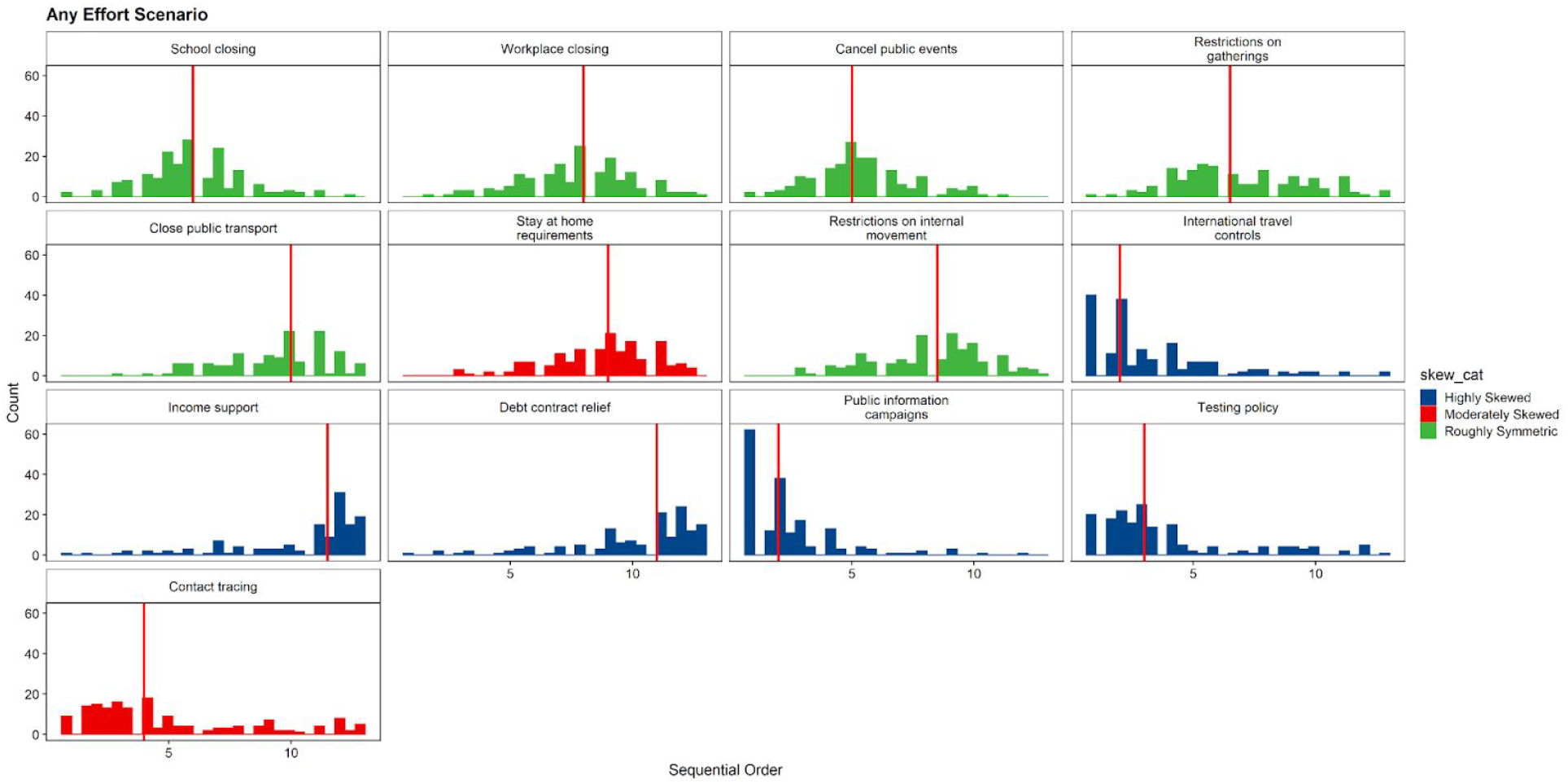
The sequential order of different NPIs under any effort scenario.

**Figure S5.**
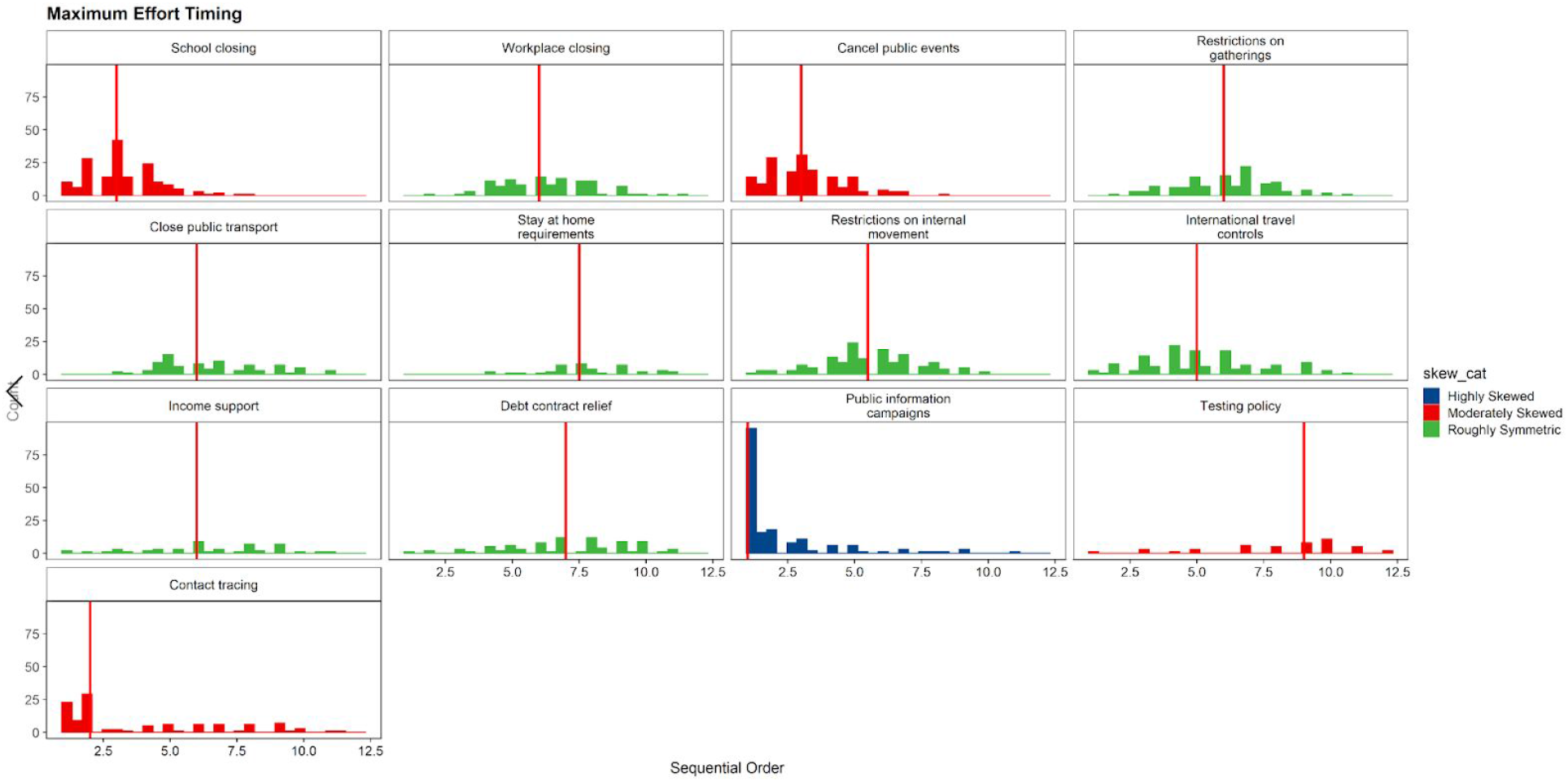
The sequential order of different NPIs under maximum effort scenario.

### A5. The *Multi-level Scenario*

Besides the *any* and *maximum efforts scenarios* described in the main text, we also investigated if intermediate levels of NPIs led to any meaningful interpretation in terms of the impacts of NPIs. In this case, original data from the Oxford Government Response Tracker is preserved, with no additional conversion. We identify two temporal clusters that cover all 13 NPIs available for the analysis.

**Figure S6.**
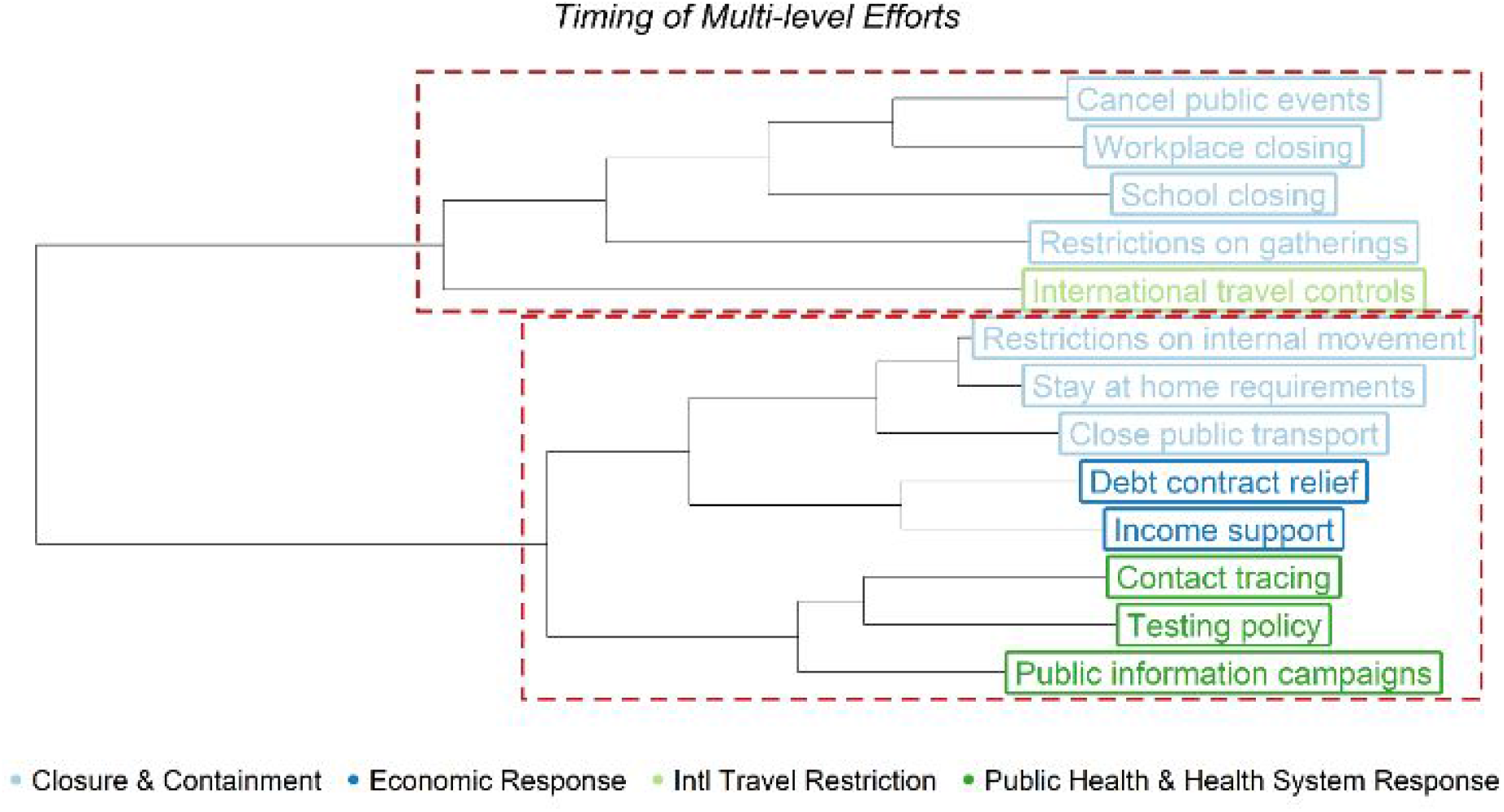
Hierarchical cluster analysis of NPIs time-series using the *multilevel scenario*. Blues and greens are used to describe the corresponding NPI groups; red boxes indicate statistically significant temporal clusters. Hierarchical clustering was carried out using Ward’s method; the statistical significance of temporal clusters identified wasobtained via bootstrapping.

**Figure S7.**
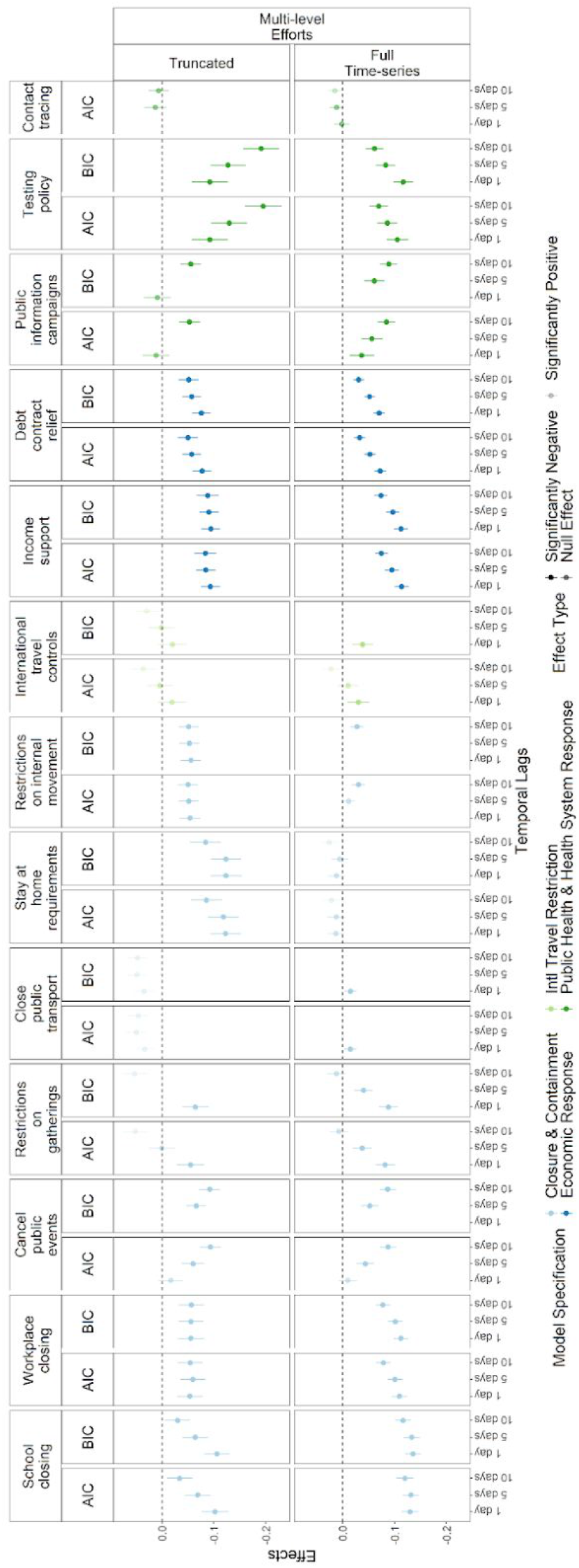
Effect sizes for each NPI from the selected models. Points and lines indicate mean and 95% confidence intervals.

Upon examining the effect sizes, we identify at least two NPIs with positive effect estimates, which indicates estimates for all NPIs are susceptible to statistical bias, preventing us from drawing reasonable conclusions. Thus, we do not include this analysis in the discussion in the main text. Current data available does not seem to allow us to examine the impacts of NPIs to this level of details.

## References

1. Kucharski AJ, Russell TW, Diamond C, Liu Y, Edmunds J, Funk S, et al. Early dynamics of transmission and control of COVID-19: a mathematical modelling study. Lancet Infect Dis. 2020 May 1;20(5):553–8.

2. Riou J, Althaus CL. Pattern of early human-to-human transmission of Wuhan 2019 novel coronavirus (2019-nCoV), December 2019 to January 2020. Eurosurveillance. 2020 Jan 30;25(4):2000058.

3. Thomas Hale, Noam Angrist, Beatriz Kira, Anna Petherick, Toby Phillips, Samuel Webster. Variation in government responses to COVID-19. BSG-WP-2020/032. Version 5.0. [Internet]. 2020 [cited 2020 May 11]. Available from: https://www.bsg.ox.ac.uk/sites/default/files/2020-05/BSG-WP-2020-032-v5.0_0.pdf

4. Barrot J-N, Grassi B, Sauvagnat J. Sectoral Effects of Social Distancing [Internet]. Rochester, NY: Social Science Research Network; 2020 Apr [cited 2020 Jun 7]. Report No.: ID 3569446. Available from: https://papers.ssrn.com/abstract=3569446

5. Quilty BJ, Diamond C, Liu Y, Gibbs H, Russell TW, Jarvis CI, et al. The effect of inter-city travel restrictions on geographical spread of COVID-19: Evidence from Wuhan, China. medRxiv. 2020 Apr;2020.04.16.20067504-2020.04.16.20067504.

6. Chinazzi M, Davis JT, Ajelli M, Gioannini C, Litvinova M, Merler S, et al. The effect of travel restrictions on the spread of the 2019 novel coronavirus (COVID-19) outbreak. Science. 2020 Apr 24;368(6489):395–400.

7. Hellewell J, Abbott S, Gimma A, Bosse NI, Jarvis CI, Russell TW, et al. Feasibility of controlling COVID-19 outbreaks by isolation of cases and contacts. Lancet Glob Health. 2020 Apr;8(4):e488–96.

8. Firth JA, Hellewell J, Klepac P, Kissler S, Kucharski AJ, Spurgin LG. Using a real-world network to model localized COVID-19 control strategies. Nat Med. 2020 Aug 7;1–7.

9. Prem K, Liu Y, Russell TW, Kucharski AJ, Eggo RM, Davies N, et al. The effect of control strategies to reduce social mixing on outcomes of the COVID-19 epidemic in Wuhan, China: a modelling study. Lancet Public Health [Internet]. 2020 Mar;0(0). Available from: http://www.ncbi.nlm.nih.gov/pubmed/32220655

10. Koo JR, Cook AR, Park M, Sun Y, Sun H, Lim JT, et al. Interventions to mitigate early spread of SARS-CoV-2 in Singapore: a modelling study. Lancet Infect Dis. 2020 Jun 1;20(6):678–88.

11. Flaxman S, Mishra S, Gandy A, Unwin HJT, Mellan TA, Coupland H, et al. Estimating the effects of non-pharmaceutical interventions on COVID-19 in Europe. Nature. 2020 Jun 8;1–8.

12. Brauner JM, Mindermann S, Sharma M, Stephenson AB, Gavenčiak T, Johnston D, et al. The effectiveness and perceived burden of nonpharmaceutical interventions against COVID-19 transmission: a modelling study with 41 countries. medRxiv. 2020 Jun 2;2020.05.28.20116129.

13. Islam N, Sharp SJ, Chowell G, Shabnam S, Kawachi I, Lacey B, et al. Physical distancing interventions and incidence of coronavirus disease 2019: natural experiment in 149 countries. BMJ [Internet]. 2020 Jul 15 [cited 2020 Aug 11];370. Available from: https://www.bmj.com/content/370/bmj.m2743

14. London School of Hygiene & Tropical Medicine. Covid-19: Global summary [Internet]. Covid-19. 2020 [cited 2020 May 11]. Available from: https://epiforecasts.io/covid/posts/global/

15. World Health Organization. Tracking public health and social measures: A global dataset. [Internet]. 2020 [cited 2020 Jun 7]. Available from: https://www.who.int/emergencies/diseases/novel-coronavirus-2019/phsm

16. World Bank. World Bank Country and Lending Groups – World Bank Data Help Desk [Internet]. 2020 [cited 2020 Jun 14]. Available from: https://datahelpdesk.worldbank.org/knowledgebase/articles/906519-world-bank-country-and-lending-groups

17. Abbott S, Hellewell J, Thompson RN, Sherratt K, Gibbs HP, Bosse NI, et al. Estimating the time-varying reproduction number of SARS-CoV-2 using national and subnational case counts. Wellcome Open Res. 2020 Jun 1;5:112.

18. Cori A, Ferguson NM, Fraser C, Cauchemez S. A New Framework and Software to Estimate Time-Varying Reproduction Numbers During Epidemics. Am J Epidemiol. 2013 Nov 1;178(9):1505–12.

19. Murtagh F, Legendre P. Ward’s Hierarchical Agglomerative Clustering Method: Which Algorithms Implement Ward’s Criterion? J Classif. 2014 Oct 1;31(3):274–95.

20. Shimodaira H, Hasegawa M. CONSEL: for assessing the confidence of phylogenetic tree selection. Bioinformatics. 2001 Dec 1;17(12):1246–7.

21. Hausman JA. Specification Tests in Econometrics. Econometrica. 1978;46(6):1251–71.

22. R Core Team. R: A language and environment for statistical computing. [Internet]. Vienna, Austria: R Foundation for Statistical Computing; 2020. Available from: http://www.R-project.org/.

23. Suzuki R, Terada Y, Shimodaira H. pvclust: Hierarchical Clustering with P-Values via Multiscale Bootstrap Resampling [Internet]. 2019. Available from: https://CRAN.R-project.org/package=pvclust

24. Croissant Y, Millo G. Panel Data Econometrics in R: The plm Package. J Stat Softw. 27(2):1–43.

25. Jackson ML, Hart GR, McCulloch DJ, Adler A, Brandstetter E, Fay K, et al. Effects of weather-related social distancing on city-scale transmission of respiratory viruses. medRxiv. 2020 Mar 3;2020.03.02.20027599.

26. Davies NG, Kucharski AJ, Eggo RM, Gimma A, Edmunds WJ, Jombart T, et al. Effects of non-pharmaceutical interventions on COVID-19 cases, deaths, and demand for hospital services in the UK: a modelling study. Lancet Public Health [Internet]. 2020 Jun 2 [cited 2020 Jun 24];0(0). Available from: https://www.thelancet.com/journals/lanpub/article/PIIS2468-2667(20)30133-X/abstract

27. Russell TW, Wu J, Clifford S, Edmunds J, Kucharski AJ, Jit M. The effect of international travel restrictions on internal spread of COVID-19. medRxiv. 2020 Jul 14;2020.07.12.20152298.

28. Clifford S, Quilty BJ, Russell TW, Liu Y, Chan Y-WD, Pearson CAB, et al. Strategies to reduce the risk of SARS-CoV-2 re-introduction from international travellers [Internet]. medRxiv. [cited 2020 Aug 10]. Available from: https://www.medrxiv.org/content/10.1101/2020.07.24.20161281v2

29. CMMID EpiForecast Core Team. [EpiForecast] Covid-19: Global summary [Internet]. EpiForecast: COVID-19. [cited 2020 May 11]. Available from: https://epiforecasts.io/covid/posts/global/

30. Stockdale JE, Doig R, Min J, Mulberry N, Wang L, Elliott LT, et al. Long time frames to detect the impact of changing COVID-19 control measures. medRxiv. 2020 Jun 16;2020.06.14.20131177.

31. Wu P, Tsang TK, Wong JY, Ng TW, Ho F, Gao H, et al. Suppressing COVID-19 transmission in Hong Kong: an observational study of the first four months. 2020;

32. Bertozzi AL, Franco E, Mohler G, Short MB, Sledge D. The challenges of modeling and forecasting the spread of COVID-19. Proc Natl Acad Sci. 2020 Jul 21;117(29):16732–8.

33. Christine Amario. Colombia’s Medellin emerges as surprise COVID-19 pioneer [Internet]. AP NEWS. 2020 [cited 2020 Jun 24]. Available from: https://apnews.com/b3f8860343323d0daeef72191b669baf

34. Linka K, Peirlinck M, Costabal FS, Kuhl E. Outbreak dynamics of COVID-19 in Europe and the effect of travel restrictions. Comput Methods Biomech Biomed Engin. 2020 May 5;0(0):1–8.

35. Lonergan M, Chalmers JD. Estimates of the ongoing need for social distancing and control measures post-“lockdown” from trajectories of COVID-19 cases and mortality. Eur Respir J [Internet]. 2020 Jul 1 [cited 2020 Aug 10];56(1). Available from: https://erj.ersjournals.com/content/56/1/2001483

36. Ghosal S, Bhattacharyya R, Majumder M. Impact of complete lockdown on total infection and death rates: A hierarchical cluster analysis. Diabetes Metab Syndr. 2020;14(4):707–11.

37. Hsiang S, Allen D, Annan-Phan S, Bell K, Bolliger I, Chong T, et al. The effect of large-scale anti-contagion policies on the COVID-19 pandemic. Nature. 2020 Jun 8;1–9.

38. Banholzer N, Weenen E van, Kratzwald B, Seeliger A, Tschernutter D, Bottrighi P, et al. Impact of non-pharmaceutical interventions on documented cases of COVID-19. medRxiv. 2020 Apr 28;2020.04.16.20062141.

39. Vaswani K. Coronavirus: The detectives racing to contain the virus in Singapore. BBC News [Internet]. 2020 Mar 19 [cited 2020 Jun 15]; Available from: https://www.bbc.com/news/world-asia-51866102

